# Spatio-temporal trends of molecularly diagnosed clinical malaria cases in The Gambia, captured across sentinel surveillance health facilities

**DOI:** 10.1101/2025.06.29.25330511

**Authors:** Eniyou Cheryll Oriero, Benjamin K. Njie, Fatoumatta Cham, Nuredin Mohammed, Brandon Nji Amambua-Ngwa, Mouhamadou Fadel Diop, Mary Oboh, Haddy Bittaye, Katty Wadda, Okebe Joseph, Annette Erhart, Umberto D’Alessandro, Serign Jawo Ceesay, Lamin Ceesay, Bolong Jobarteh, Abdoulie Jarju, Ebba Secka, Alhagie Sankareh, Jeandarc Jarju, Lamin Sanneh, Sheriffo Jagne, Olimatou Kolley, Momodou Kalleh, Ballah Gibba, Wandifa Samateh, Alfred Amambua-Ngwa

## Abstract

**Objectives:** Malaria transmission is becoming more heterogeneous across sub-Saharan Africa, and elimination efforts in low-transmission countries are under threat of local or imported antimalarial resistance. The Genomic Surveillance of Malaria (GSM) project established a hub in The Gambia to support malaria elimination efforts through enhanced parasite surveillance as interventions are intensified. We report trends in clinical malaria cases at sentinel sites confirmed by molecular diagnosis over a 4-year period (2019 to 2022).

**Methods:** A total of 10,052 study participants from 383 villages were recruited at 23 health facilities in the six regions of the country. Parasite positivity rates were compared and complexity of infection (COI) was assessed to distinguish spatial and temporal epidemiologically relevant differences.

**Results:** Across the years and all regions, PCR-positive proportions of suspected malaria cases ranged from 9.4% - 73%, with significantly more clinical cases detected in the semi-urban western sites over time. Approximately 12.9% of overall infections detected by PCR were missed by RDT across all regions and there was an upward trend of single-strain *P. falciparum* infections over the study period.

**Conclusion:** Integrating molecular approaches in routine malaria surveillance provides high-resolution data for prevalence stratification and customized interventions for targeted local malaria elimination strategies.

## INTRODUCTION

An estimated 263 million malaria cases were reported globally in 2023, and 249 million in 2022, a steady increase from 245 million cases reported in 2020. Most of the increase was observed in the World Health Organization (WHO) African Region [1, 2]. A major strategy in malaria control is the effective confirmation and treatment of clinical malaria detected through passive surveillance integrated into the public health infrastructure. Elimination programs leverage on sustaining controlled low-endemic malaria to interrupt transmission and prevent its re-establishment [3]. Recent emergence of partial Artemisinin resistance in *Plasmodium falciparum* in sub-Saharan African countries poses a significant threat to case management and the global effort to reduce the disease burden [4, 5]. A review of the potential human and economic costs of widespread resistance to Artemisinin combination therapies (ACTs) determined a hypothetical scenario that resulted in yearly excess of millions in treatment failures, excess deaths, and management costs, including estimated costs associated with changes in treatment policy [6]. Thus, the deployment of sensitive molecular and genomic technologies for malaria surveillance in low-transmission settings has the potential to improve monitoring of trends in malaria transmission and markers of treatment responses, which could potentially affect efficacy of chemotherapeutic and chemopreventive interventions used by National Malaria Programmes (NMPs).

The NMPs across Africa are faced with multiple and significant challenges, including the efficacy of frontline treatments as well as available alternatives, identification of transmission hubs, importation of cases, possible routes of spread, etc., using mostly clinical and epidemiological data available from routine clinical reporting and efficacy studies [7]. One of the pillars in the strategic framework by the WHO to accelerate progress toward malaria elimination is to transform malaria surveillance into a key intervention [8]. Therapeutic efficacy data can be supplemented by surveillance of antimalarial drug resistance, which is required for early detection of parasite adaptation to drugs and changing patterns of susceptibility, to enable timely revisions of national and global antimalarial drug policies [6]. In addition, the significant spatial heterogeneity and seasonal fluctuations in the distribution of malaria and asymptomatic carriage in areas where transmission has become low or moderate highlights the importance of increased surveillance to identify the most susceptible demographic groups and hotspots for targeted interventions [8]. As malaria interventions are upscaled, it is expected that clinical cases and asymptomatic infections at low parasite densities will become common [3].

In low malaria transmission areas, strategies such as reactive afebrile case detection from index clinical infections allow for effective detection of clusters of infections and transmission hotspots. Layering molecular analysis on these approaches will further enable detection of sources, sinks, and levels of relatedness of low-grade afebrile malaria infections [9]. In addition to routine intervention, intensive case-and foci-based surveillance to identify and reduce reservoirs of infections, and prevent transmission from imported cases are required as endemic regions advance to malaria pre-elimination [10]. These approaches will benefit from more sensitive case detection and spatial analysis to identify hotspots and inform focal interventions. Although malaria transmission is overall low across the Gambia, it remains heterogeneous across health districts, with moderate transmission sustained in the eastern region bordering the most endemic regions of neighbouring Senegal. However, the National Malaria Control Program (NMCP) strategizing towards pre-elimination, targets selected health districts in which infection prevalence is at or lower than 0.1% [11]. To support this ambition, this study reports the rate of molecularly detected infections from suspected clinical malaria cases in The Gambia between 2019 to 2022, where RDT is used for routine diagnosis and treatment. This data supports a high-resolution, nationwide, proportional stratification and malaria molecular surveillance program for antimalarial resistance and population dynamics for determination of parasite transmission.

## METHODS

### Study sites and populations prior to the genomic surveillance program

Clinical malaria data was extracted from three earlier studies conducted by the Medical Research Council Unit The Gambia (MRCG) spanning 13 years (2005 – 2018). These were analyzed to estimate clinical malaria case recruitment before 2019. The studies were conducted in health facilities from two regions in the country, one in the coastal region where malaria decline had previously been reported (Brikama); and the other in the eastern part of the country with significant residual malaria transmission (Basse), summarized in the **Supplemental Table 1**. The studies employed passive case detection of uncomplicated malaria infections, including severe malaria cases in one of the studies.

### Network of sentinel health facilities for malaria genomic surveillance in The Gambia

In consultation with the Gambian NMCP, health facilities were chosen following a review of routine reported clinical malaria prevalence data and planned prioritization of interventions implemented in different regions of the country. Health facilities retained spanned the six administrative regions of the country. The study period from 2019 to 2022 coincided with the Covid19 pandemic, during which the lock-down and other restrictions interrupted regular delivery of some health services. Across the study period, participants were recruited from 23 health facilities – 4 in the Western region, 2 in the North Bank region, 3 in the Lower River region, 8 in Central River region, and 6 in the Upper River region (**Figure 1**). Specific interventions implemented during the study period included seasonal malaria chemoprevention (SMC) administered to children under 5 years of age in selected health districts in the eastern regions of the country (Central and Upper River Regions). Scientific review and Ethics approval for the study were obtained from the MRCG’s Scientific Coordinating Committee and the Joint Gambian Government/Medical Research Council Unit The Gambia Ethics Committee (Ref: SCC 1629). The staff in each study health facility were trained on consenting and blood sample collection at the beginning of each malaria transmission season. For engagement, meetings were held with the Regional Health Directors (RHDs) and the NMCP before and after each malaria transmission season to review progress and challenges to the study timelines and objectives.

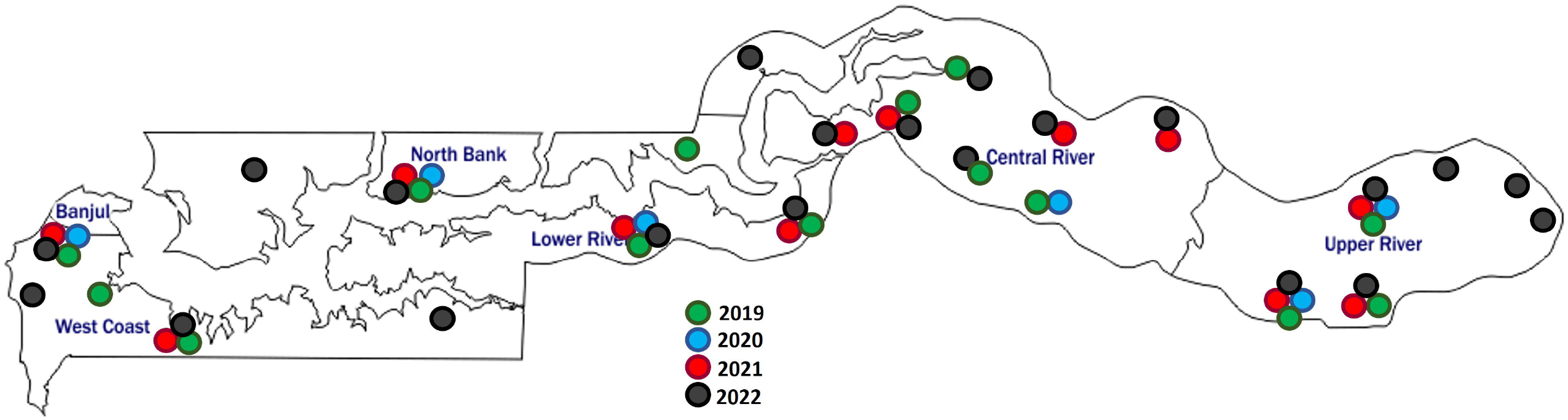

### Participant recruitment and sample collection

All individuals who presented to the study health facilities with suspected malaria fever or were referred for malaria testing were eligible to participate. Study participants were recruited upon provision of written informed consent as well as an assent for eligible children, after which relevant metadata such as age, gender, village, and prior antimalarial use were also obtained. Each year, enrolment and sample collection was done between September and December; during the malaria transmission season. Sample collection kits were provided to each health facility, including lancets and swabs for finger pricking, and dried blood spot (DBS) cards (Whatman 3M filter paper) for each participant with unique sample ID barcode labels, (**Supplemental Figure 1**). A single finger prick was used for both rapid diagnostic test (RDT) and DBS collection, according to the Malaria Genetic Epidemiology Network (MalariaGEN) SpotMalaria protocol [12]. Four blood spots from each participant were made on the DBS cards provided. The RHDs provided oversight and supervision of the health facility surveys while the MRCG team provided training of health facility staff for sample collection, along with periodic monitoring of field activities.

### Molecular detection of malaria parasites

DNA was extracted from three 6mm blood spots punched using a manual paper puncher. The paper puncher was decontaminated in between samples with 10% bleach solution and then rinsed with Millipore Alpha Q water. DNA was extracted using the automated QIAcube HT according to manufacturers’ protocols (Qiagen GmbH, Hilden, Germany). The presence of malaria parasites in the samples was detected on the Biorad CFX96 real-time PCR machine using an ultra-sensitive qPCR protocol that targets the *var* gene acidic terminal sequence (*var*ATS) [13]. A threshold Cq ≤ 40 was used to determine positive samples, and a no-template negative control was included in each run as a quality check for operator-related cross-contamination. Quality control was done for all assay steps from DBS punching and DNA extraction to the nucleic acid amplification assays.

### Data collection and analysis

The data from the case report forms collected in each health facility were entered onto a REDCap electronic data capture tool [14, 15]. Study data extracted from REDCap were analyzed using GraphPad Prism Version 10.2.1 (395) and R statistical software v4.2.2. Global Positioning System (GPS) coordinates for the health facilities and participants’ villages were used to generate visual-spatial maps using QGIS v3.34. The proportion of PCR-positive cases was determined from the suspected malaria cases recruited in each health facility and compared against RDT results, where both results were available. A Bayesian approach (THE REAL McCOIL) using Markov chain Monte Carlo methods to simultaneously estimate allele frequency and complexity of infection (COI), which is an indication of transmission intensity, was determined to distinguish spatial and temporal epidemiologically relevant differences in COI [16]. A spatial display of the reported villages of the study participants was analyzed.

## RESULTS

### Baseline clinical malaria data (2005 – 2018)

Three retrospective studies provided the baseline estimates of clinical malaria recruitment before 2019. Characteristics of participants recruited in these clinical studies show an increase in the mean age over time, from 3.9 in 2005 to 19.0 in 2018, and a marked decline in the parasite density (%parasitemia) (**Figure 2a**), consistent with the decline in malaria transmission observed in The Gambia. A slight increase was also observed in the hemoglobin levels, from 8.5g/dl to 11.11g/dl within 5 years (**Figure 2b**), possibly due to the increasing mean age of individuals with malaria over time.

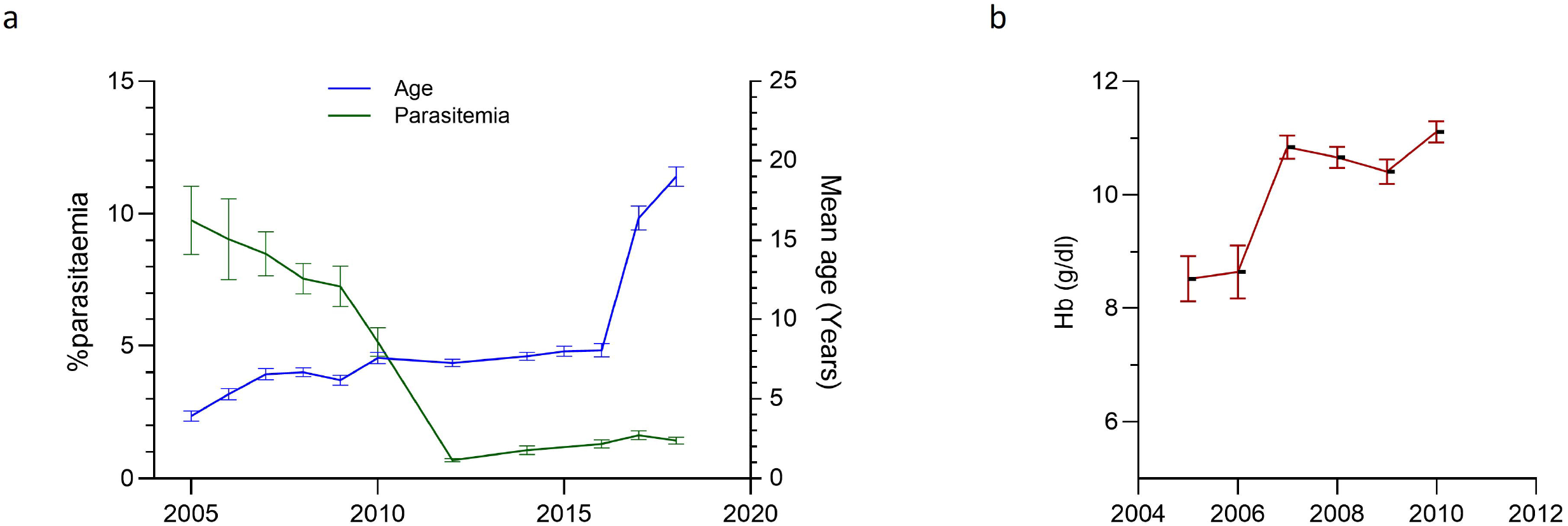

### Demography of the study population and heterogeneity of molecularly diagnosed malaria across the Gambia

A total of 10,052 participants were recruited across the four years of the GSM, with similar proportions of male and female participants, and age categories. A significantly higher number of participants were above 15 years old in all four years of the study period (**Supplemental Table 2**).

Individuals with suspected malaria sampled at the sentinel health facilities came from a median of 311 (range 238 – 383) villages annually, while PCR-positive cases were recorded in individuals from a median of 193 (range 150 – 253) villages (**Supplemental Table 3**). Overall, the villages of PCR-positive cases observed each year were similar, though notable shifts were seen in Western Region 2 (67.7% to 80.9%), and Upper River Region (86.7% to 66.7%).

For individuals with both PCR and RDT results, positivity rates were compared at health facility and regional levels, and this consistently detected significantly more *P. falciparum* infections (p < 0.05) by molecular methods (**Table 1**). RDT detected approx. 69.9% of all possible positive malaria infections (obtained by combining all positive cases by either PCR or RDT), PCR detected approx. 86.3% and both methods agreed on 56.3%.

**Table 1:** Comparison of malaria positivity by PCR and RDT for study participants per health facility.

| S/N | Health Facility | Region | Total | RDT+ (%) | PCR+ (%) | RDT+PCR+ (%) | RDT+ or PCR+ (%) | RDT-PCR+ (%) | RDT+PCR- (%) | RDT-PCR- (%) |
| --- | --- | --- | --- | --- | --- | --- | --- | --- | --- | --- |
| 1 | Brikamaba | CRR | 386 | 50 (12.95) | 71 (18.39) | 31 (8.03) | 90 (23.32) | 40 (10.36) | 19 (4.92) | 296 (76.68) |
| 2 | Kuntaur | CRR | 229 | 57 (24.89) | 89 (38.86) | 53 (23.14) | 93 (40.61) | 36 (15.72) | 4 (1.75) | 136 (59.39) |
| 3 | Soma | LRR | 1165 | 99 (8.49) | 302 (25.92) | 82 (7.04) | 319 (27.38) | 220 (18.88) | 17 (1.46) | 846 (72.62) |
| 4 | Pakaliba | LRR | 583 | 163 (27.96) | 201 (34.48) | 129 (22.13) | 235 (40.31) | 72 (12.35) | 34 (5.83) | 348 (59.69) |
| 5 | Kerewan | NBR | 1058 | 130 (12.29) | 177 (16.73) | 100 (9.45) | 207 (19.57) | 77 (7.28) | 30 (2.84) | 851 (80.43) |
| 6 | Sabi | URR | 253 | 134 (52.96) | 154 (60.87) | 121 (47.83) | 167 (66.01) | 33 (13.04) | 13 (5.13) | 86 (33.99) |
| 7 | Yorobawol | URR | 917 | 362 (39.48) | 419 (45.69) | 294 (32.06) | 487 (53.11) | 125 (13.63) | 68 (7.42) | 430 (46.89) |
| 8 | Sotuma | URR | 465 | 300 (64.52) | 316 (67.96) | 256 (55.05) | 360 (77.42) | 60 (12.90) | 44 (9.46) | 105 (22.58) |
| 9 | Fajikunda | WR1 | 968 | 307 (31.71) | 401 (41.43) | 263 (27.17) | 445 (45.97) | 138 (14.26) | 44 (4.54) | 523 (54.03) |
| 10 | Pirang | WR2 | 151 | 34 (22.52) | 39 (25.83) | 22 (14.57) | 51 (33.77) | 17 (11.26) | 12 (7.95) | 100 (66.23) |
| 11 | Kafuta | WR2 | 809 | 322 (39.80) | 347 (42.89) | 273 (33.75) | 396 (48.95) | 74 (9.15) | 49 (6.06) | 413 (51.05) |
| 12 | Bansang | CRR | 549 | 214 (38.98) | 242 (44.08) | 176 (32.06) | 280 (51.00) | 66 (12.02) | 38 (6.92) | 269 (48.99) |
| 13 | Ngeynen Sanjal | NBR | 21 | 1 (4.76) | 3 (14.29) | 1 (4.76) | 3 (14.29) | 2 (9.52) | 0 | 18 (85.71) |
| 14 | Sami Karantaba | CRR | 161 | 26 (16.14) | 50 (31.06) | 25 (15.52) | 51 (31.68) | 25 (15.53) | 1 (0.62) | 110 (68.32) |
| 15 | Janjangbureh | CRR | 229 | 85 (37.12) | 108 (47.16) | 64 (27.94) | 129 (56.33) | 44 (19.21) | 21 (9.17) | 100 (43.67) |
| 16 | Kudang | CRR | 94 | 5 (5.32) | 16 (17.02) | 3 (3.19) | 18 (19.15) | 13 (13.83) | 2 (2.13) | 76 (80.85) |
| 17 | Keneba | LRR | 93 | 51 (54.84) | 48 (51.61) | 40 (43.01) | 59 (63.44) | 8 (8.60) | 11 (11.83) | 34 (36.56) |
| 18 | Kaur | CRR | 89 | 61 (68.54) | 59 (66.29) | 48 (53.93) | 72 (80.89) | 11 (12.35) | 13 (14.61) | 17 (19.10) |
| 19 | Kuntaya | NBR | 192 | 81 (42.19) | 80 (41.67) | 62 (32.29) | 99 (51.56) | 18 (9.38) | 19 (9.89) | 93 (48.44) |
| 20 | Baja Kunda | URR | 111 | 36 (32.43) | 44 (39.64) | 17 (15.31) | 63 (56.75) | 27 (24.32) | 19 (17.12) | 48 (43.24) |
| 21 | Fatoto | URR | 245 | 82 (33.47) | 68 (27.75) | 39 (15.92) | 111 (45.31) | 29 (11.84) | 43 (17.55) | 134 (54.69) |
| 22 | Koina | URR | 138 | 33 (23.91) | 52 (37.68) | 29 (21.01) | 56 (40.58) | 23 (16.67) | 4 (2.89) | 82 (59.42) |
| 23 | Tanji | WR2 | 289 | 138 (47.75) | 135 (46.71) | 102 (35.29) | 171 (59.17) | 33 (11.42) | 36 (12.46) | 118 (40.83) |
|  | Total |  | 9195 | 2771 (30.14) | 3421 (37.21) | 2230 (24.25) | 3962 (43.09) | 1191 (12.95) | 541 (5.88) | 5233 (56.91) |

Overall, approximately 12.9% of infections detected by PCR were missed by RDT across all regions (**Figure 3a**), with proportions observed in the health facilities ranging from 7.3% (Kerewan – NBR) to 24.3% (Baja Kunda – URR). On the other hand, approximately 5.8% who did not have parasites detected by PCR were reported and treated as RDT-positive.

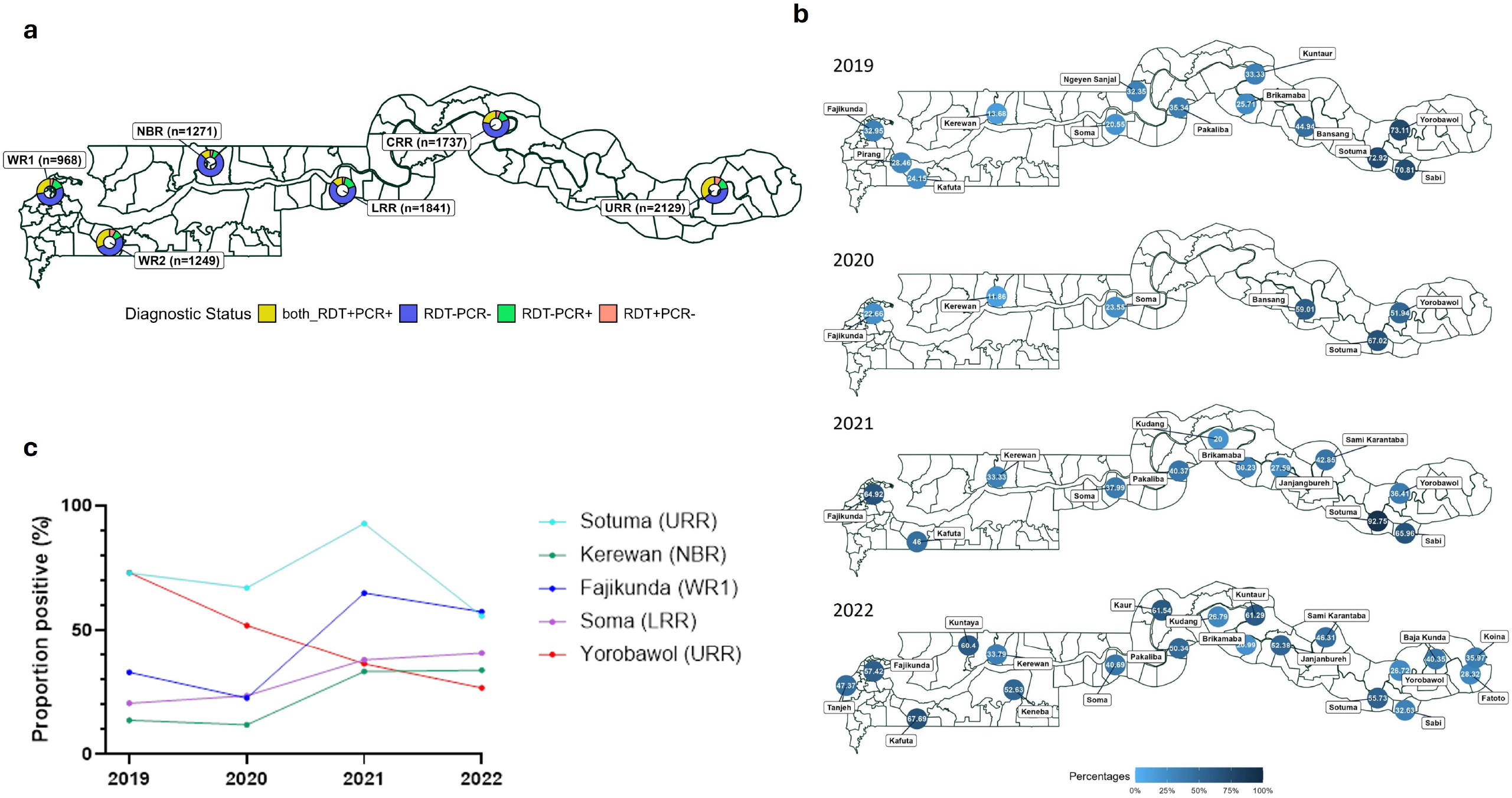

A comparison of the proportion of positive cases per health facility showed heterogeneity of malaria infections and temporal trends across the country. Specifically, the proportion of positive cases in the Eastern (inland, rural) part of the country with moderate malaria transmission, reduced in 2022 compared to the 2019 baseline, considering the sites of Yorobawol, Sotuma, and Sabi as examples (**Figure 3b**). A reverse trend was observed in the Western (coastal, urban) parts of the country which historically, has reported lower malaria prevalence (e.g. Fajikunda Kafuta, etc.). Exploring this trend further in five health facilities with complete data across the study period (**Figure 3c**), showed a decline in the two health facilities in the Upper River Region (Yorobawol and Sotuma), a marked increase in the Western Region (Fajikunda), a modest but steady increase in the North Bank (Kerewan) and the Lower River (Soma) administrative regions, highlighting possible stochasticity expected in low transmission settings

The complexity of infection (COI) analysis showed predominantly single-strain infections of *P. falciparum* across all regions of the country, ranging from 52 – 100% (**Figure 4**). An overall upward trend of single-strain infections was observed between 2019 and 2022, with a significant reduction in multiple-strain infections - from 26.29% to 11.74% (P < 0.05).

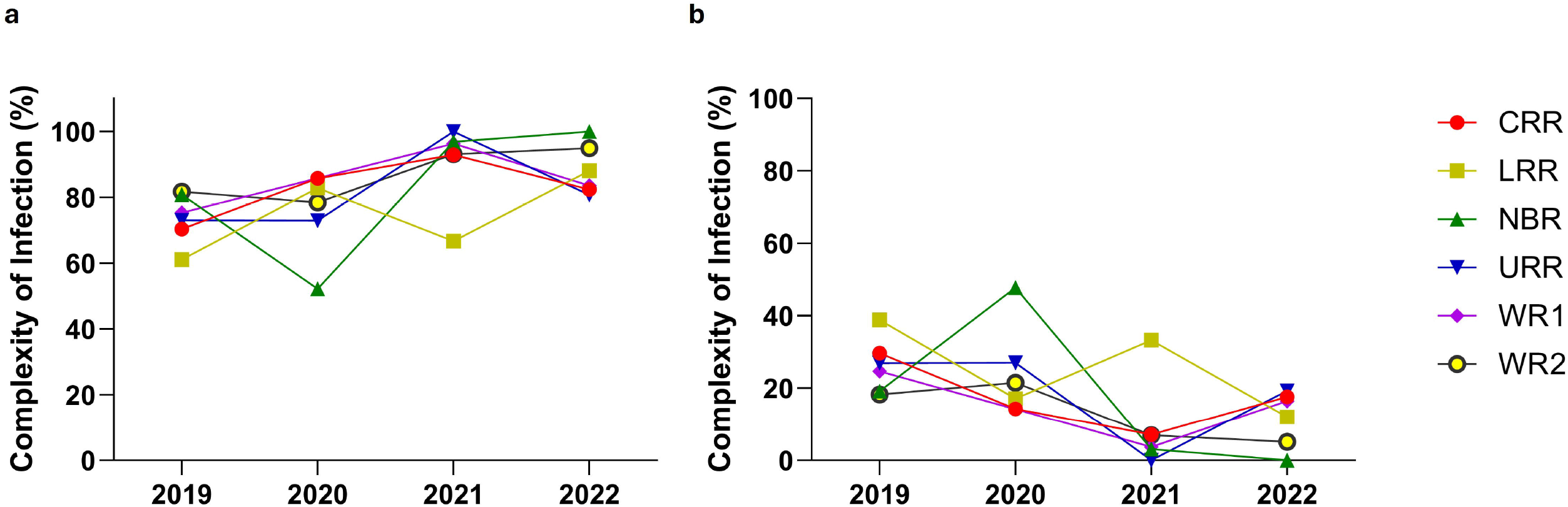

The spatial distribution and coverage of villages across the country increased from 2019 to 2022 as more health facilities were added to the study post-COVID-19. However, the density of suspecte d malaria cases increased by proximity to a study health facility across all years (**Figure 5**), with 80-90% of the villages being within 50 Km from the health facility that the participants reported.

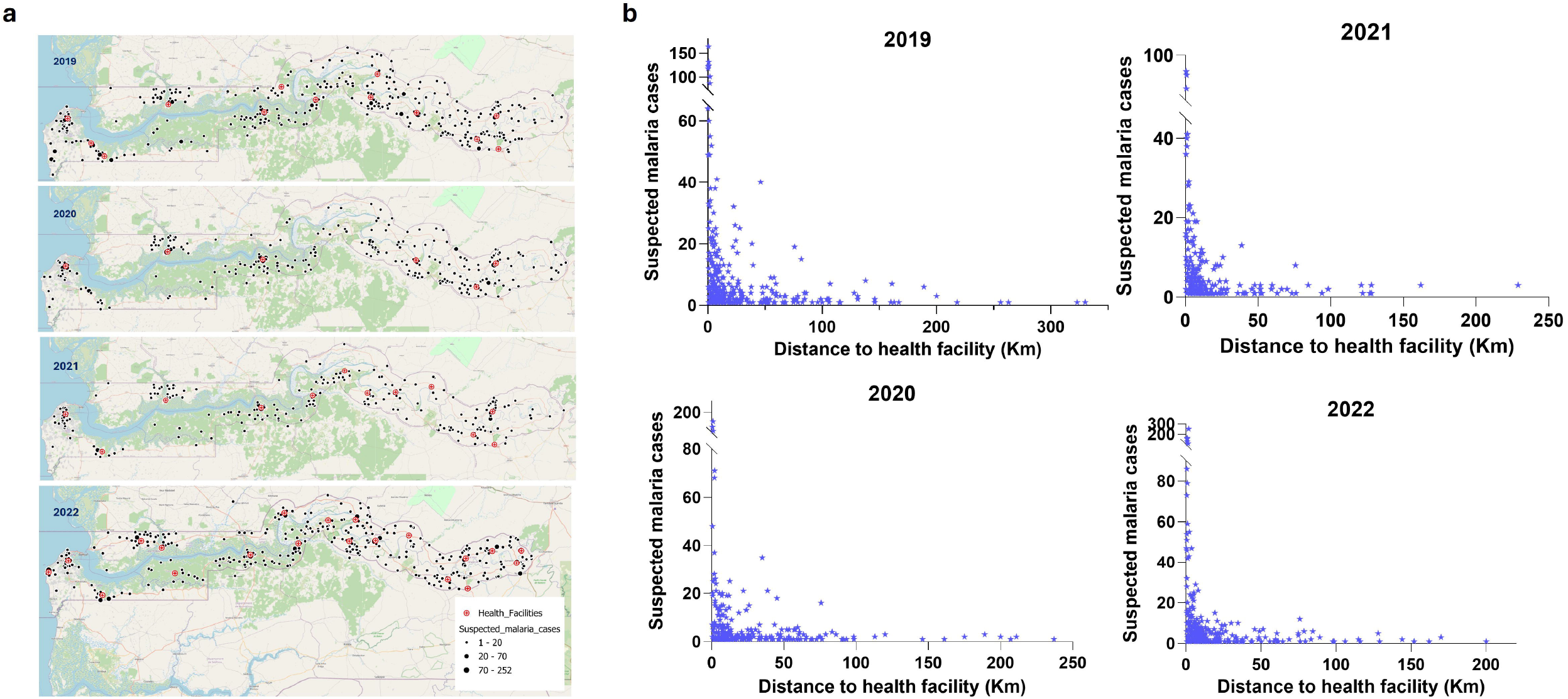

## DISCUSSION

We present the malaria positivity rates of consenting individuals who attended the sentinel surveillance health facilities with suspected malaria disease, detected by the sensitive molecular test, and compared against the WHO-recommended malaria RDT (where data was available). Malaria prevalence had significantly declined and the NMCPs strategy envisions pre-elimination. This strategy is in line with the low parasitemia observed in the baseline survey (2012 - 2018), which was consistent with the previously reported decline in malaria transmission in The Gambia [17, 18]. Low parasitemia levels pose peculiar challenges to accurate detection with the currently recommended routine diagnostic tools – microscopy and RDT. Thus molecular detection of cross-sectional samples from endemic clinical populations could provide more accurate estimates of malaria parasite infection prevalence at sentinel health facilities. Currently, the NMCPs rely on records of RDT-diagnosed malaria for reporting trends and defining layers of country stratification and intervention strategies. Here, we found missing RDT results from some health facilities, which could mostly be attributed to the paper-based recording system still in practice in Gambian Health facilities. By collecting barcoded RDT cassettes, case records, and dried blood spots from each participant, more complete records were subsequently obtained. Hence, sentinel surveillance of malaria towards elimination may benefit from this sort of advanced case-recording method.

The proportion of malaria cases missed by RDT (approximately 13% overall) was not negligible and could increasingly become more significant as countries progress towards malaria elimination. The wide heterogeneity of low-density infections observed in this low transmission setting (ranging from 7.3% to 24.3%), supports the narrative that a one-size-fits-all approach may not be an ideal strategy for malaria elimination interventions. This is also reflected in the higher proportions of RDT-positive cases (up to 17.5%) that were PCR-negative, possibly due to persistent antigen circulation from a recent malaria infection. Unfortunately, the history of malaria infection was estimated from information obtained on recent antimalarial use prior to being recruited into the study, which was inadequate to make substantial conclusions.

Overall, we show low complexity of infections across all regions of the country, consistent with the low transmission status of The Gambia. Estimates of complex or multiple-strain *P. falciparum* infection prevalence and population-average COIs correlate with transmission intensity and potential indicators of disease control efforts due to the ubiquitous nature of parasite co-transmission and superinfection which scales with the entomological inoculation rate [19]. Sentinel surveillance of malaria by NMCPs collect binary data from RDT-based malaria diagnosis. However, refined planning of malaria elimination may need higher granularity such as levels of parasiteamia, complexity of infection, and their distribution in demographic groups. This is even more important with the introduction or plans to implement new interventions such as vaccines. Such key datasets, which can be derived from research need to be harmonized with public health information, as demonstrated here.

We showed significant spatial heterogeneity in the subset of infections (consenting study participants) analyzed per health facility, which also varied over time. In line with annual reports, malaria in 2019 was more prevalent in the eastern end of the country, which is furthest into Senegal. These regions border the southeast regions of Senegal which have the highest prevalence and transmission of malaria. As such, epidemiological and genomic surveillance of malaria will benefit from cross-border collaboration. The Genomic Surveillance of Malaria (GSM) Project therefore has established collaboration for mapping genetic markers of parasites at sites in Senegal, that border The Gambia, while the Gambian NMCP are working with their counterparts in Senegal to synchronise interventions (e.g LLINs distribution). In addition, spatiotemporal presentation of clinical malaria cases in maps can be explored by NMPs to monitor the impact of interventions across health districts. We also observed temporal change in the spatial variance of infection, with prevalence becoming higher in the western semi-urban coastal regions in 2022, a reversal of the pattern seen in 2019. The survey period spanned the period of emergence and spread of SARS-CoV-2, and the COVID-19 pandemic. COVID-19 interrupted healthcare provision and interventions such as lockdowns confined individuals in households, which we have previously shown to be the core unit of transmission of malaria [20]. Therefore, the expansion of malaria in western Gambia could partly be due to COVID-19, warranting genetic analysis to show that infections detected were indeed locally transmitted and if those in households were all related. There were also recent spike in severe malaria cases reported in a tertiary hospital in the urban (coastal) region of the country from the peak of the pandemic [21]. Taken together, the re-bound of malaria in previously low transmission regions calls for reassessment of the malaria elimination strategy by the Gambian NMCP. The program had been implementing seasonal malaria prevention alongside other interventions mainly in the East of the country where transmission had been moderate. National re-stratification based on more recent surveys with additional sensitive diagnosis as reported here should provide the needed evidence for data-driven decision-making about intervention strategies and policies.

The health facilities included here for sentinel surveillance of malaria were chosen in consultation with the NMCP and we observed that most of the cases captured were close to the health facility, an effect of proximity previously reported in other public health surveillance programs [22]. This is a limitation that could be remedied by community-based surveillance, which unfortunately is more costly and harder to fund. Alternatively, subsequent surveys could include informal and private health facilities such as drug stores and private clinical labs, which provide healthcare services at community levels, especially for febrile illnesses like malaria.

Molecular tools for malaria parasite detection will become increasingly important as transmission is driven down and clinical malaria cases result from infections with lower parasite densities. Integrating molecular surveillance data with genomic data for different public health use cases such as monitoring the impact of interventions e.g. antimalarial resistance and monitoring malaria transmission, provides the best resolution and serves as early warning signs to inform policy on these use cases.

## Data Availability

All data produced in the present study are available upon reasonable request to the authors

## ACKNOWLEDGMENT

We acknowledge all the participants who contributed to the success of this study and all past and current members of the Malaria Population Biology Group at MRCG.

## FINANCIAL SUPPORT

This research was funded by the NIHR (134717) using UK international development funding from the UK Government to support global health research. The views expressed in this publication are those of the author(s) and not necessarily those of the NIHR or the UK government. Additional support to the Malaria Population Biology Group at MRCG for molecular epidemiology of malaria was received from the PAMGEN project, an H3Africa consortium funded by Science for Africa Foundation (H3AFull/17/008). and the EGSAT senior fellowship plus to Prof Amambua funded by the EDCTP (TMA2019SFP-2843)

## ROLE OF THE FUNDING SOURCE

The study’s funder had no role in study design, data collection, data analysis and interpretation, or report writing.

## CONFLICT OF INTEREST

The authors have no conflict of interest.

## Notes

### Competing Interest Statement

The authors have declared no competing interest.

### Author Declarations

The Joint Gambian Government/Medical Research Council Unit The Gambia Ethics Committee gave approval for this work

### Summary of Updates

The funding statement was updated at the request of the funders. The author list was also updated.

